# Safe in my heart: resting heart rate variability longitudinally predicts emotion regulation, worry and sense of safeness during COVID-19 lockdown

**DOI:** 10.1101/2021.06.17.21259071

**Authors:** Elena Makovac, Luca Carnevali, Sonia Hernandez-Medina, Andrea Sgoifo, Nicola Petrocchi, Cristina Ottaviani

## Abstract

Due to its ability to reflect the capacity to engage in context-appropriate responses, tonic heart rate variability (HRV) is considered a putative biomarker of stress resilience. However, most studies are cross-sectional, precluding causal inferences. The high levels of uncertainty and fear at a global level that characterize the COVID-19 pandemic offer a unique opportunity to investigate the longitudinal role of HRV in stress resilience. The present study examined whether HRV, measured about 2 years earlier (Time 0), could predict emotion regulation strategies and daily affect in healthy adults during the May 2020 lockdown (Time 1). Moreover, we evaluated the association between HRV measures, emotion regulation strategies, subjective perception of COVID-19 risk, and self-reported depressive symptoms at Time 1. Higher tonic HRV at Time 0 resulted a significant predictor of a stronger engagement in more functional emotion regulation strategies, as well as of higher daily feelings of safeness and reduced daily worry at Time 1. Moreover, depressive symptoms negatively correlated with HRV and positively correlated with the subjective perception of COVID-19 risk at Time 1. Current data support the view that HRV might be not only a marker but also a precursor of resilience under stressful times.

## INTRODUCTION

On March 11, 2020, the COVID-19 outbreak was declared a global pandemic (1), and almost all countries in the world have introduced national lockdowns. Psychosocial efforts to resist the lockdown situation first and overcome the negative consequences of the pandemic have caused mental health problems and continue to challenge vulnerable individuals (2-4). Indeed, the identification of risk, vulnerability and protective factors for mental health has been recognised as a research priority (5-7).

Tonic vagally-mediated heart rate variability (vmHRV), an index of parasympathetic cardiac modulation, has been indicated as a biomarker of stress resilience, as it reflects the ability to effectively regulate emotions in a changing environment (8-10). Unfortunately, the majority of such investigations is cross-sectional and longitudinal studies on the putative role of vmHRV as a predictor of stress resilience are sparse. For example, preliminary studies have shown that lower HRV before or in the aftermath of a traumatic experience predicts the development of PTSD symptoms in adults (11). To the best of our knowledge, the association between vmHRV and COVID-19-related stress was examined only in one study, in which changes in HRV parameters during and after the lockdown period appeared to be in line with subjective well-being (12). Moreover, in a study conducted on U.S. residents between March and May 2020, self-reported autonomic reactivity measured by The Body Perception Questionnaire Short Form (13) mediated the relation between past traumatic experiences and mental health indices during the COVID-19 pandemic (14). However, a recent study in adolescents found that susceptibility to mental health difficulties associated with COVID-19 stress was predicted by higher resting vmHRV assessed 4 years earlier (15), which is inconsistent with cross-sectional and longitudinal findings in adults.

The present investigation aimed at extending previous findings, with the hypothesis that higher resting vmHRV, assessed ∼22 months earlier, would predict the use of more functional emotion regulation strategies and daily affective states in healthy adults during COVID-19-related forced lockdown. Furthermore, we tested the cross-sectional relationship between vmHRV, depressive symptomatology, and subjective perception of COVID-19 risk during the lockdown.

## METHODS

### Procedure

One-hundred and seventy-two healthy individuals, whose resting vmHRV was assessed between May 2018 and October 2019 (Time 0; T0) during participation in previous studies (16-19), were recruited. Specific information about vmHRV assessment can be found in the original studies (16-19). After the exclusion of 2 participants with current diagnosis of medical disorders, a total of 69 individuals were considered for this investigation, which includes data collected during national lockdowns in Italy and United Kingdom from May 1 to May 31, 2020 (Time 1; T1) (ClinicalTrials.gov identification code: NCT04382560). Participants provided informed consent, and the study was approved by the IRBs of King’s College London (LRS-19/20-18429:COVID-19) and Sapienza University of Rome (Prot. 0000653).

Initially, participants filled out online questionnaires including (i) the Perceived Coronavirus Risk scale (PCRS;(20)), which measures the subjective perception of COVID-19 risk; (ii) the Center for Epidemiological Studies Depression Scale (CES-D;(21)), which assesses self-reported depressive symptoms experienced in the previous week; and (iii) the 10-item Emotion Regulation Questionnaire (ERQ; (22)), which measures the habitual use of several emotion regulation strategies. Among them, the present study focused on cognitive reappraisal and emotional suppression as examples of functional and dysfunctional strategies, respectively. Next, participants were asked to download a free-avilable smartphone application (https://ecg4everybody.com/) and instructed to record vmHRV using finger tip photoplethysmography under resting conditions. Specifically, they were asked to refrain from drinking coffee and alcholic and energizing drinks, eating a meal, smoking, and physical exercising for at least two hours prior to the recording, similar to the T0 assessment. Subsequently, participants were asked to rate their current levels of feeling anxious, sad, worried, optimistic, and safe on 5-point Likert scales every 2 hours during wake for 2 consectuive days, using electronic diaries delivered by Qualtrics (23).

### Analyses

The root mean square of successive differences between normal heartbeats was considered as an index of vmHRV (24). Pearson’s correlations and *t*-tests were performed to investigate the influence of age, sex, lifestyle factors (e.g., smoking) and recording procedures on vmHRV values at T0 and identify the potential covariates to include in the subsequent regression models. First, we evaluated whether vmHRV values at T0 would predict reappraisal and suppression ERQ scores at T1, by applying linear regressions. Next, we tested whether vmHRV values at T0 would predict momentary mood ratings at T1, by applying random effects regression models with maximum likelihood estimation. Lastly, Pearson’s correlations were computed between vmHRV, CESD, PCRS, and ERQ scores at T1, controlling for the effects of age and sex.

## RESULTS

The final sample size, excluding incomplete responses (*n* = 3), was *n* = 66 (Table 1). No significant associations emerged between baseline participants’ demographic, anthropometric, and lifestyle characteristics (e.g., age, sex, BMI, smoking status) and vmHRV at T0. Therefore, only the months elapsed between T0 and T1 were controlled for in the regression models. As shown in Table 2, vmHRV at T0 positively and negatively predicted the use of cognitive reappraisal and emotional suppression at T1, respectively. As to momentary mood, the models having safe and worried at T1 as outcomes yielded a significant role of vmHRV at T0 as predictor (Table 3), whereas no significant associations emerged for anxious, sad, and optimistic.

**Table 1.** Participants’ characteristics (*n* = 66).

|  |  |
| --- | --- |
| Sex | Females 63.6% |
| Age (years) at Time 0 | $24.0 \pm 5.9$ |
| BMI ($\text{kg}/\text{m}^2$) at Time 0 | $23.2 \pm 3.8$ |
| Smoking status at Time 0 | Smokers 55% |
| HR (bpm) at Time 0 | $73.1 \pm 13.0$ |
| lnvmHRV at Time 0 | $3.73 \pm 0.59$ |
| Months elapsed between Time 0 and Time 1 | $21.5 \pm 2.4$ |
| ERQ score (reappraisal subscale) at Time 1 | $28.3 \pm 8.4$ |
| ERQ score (suppression subscale) at Time 1 | $13.9 \pm 4.6$ |
| CESD score at Time 1 | $15.3 \pm 4.7$ |
| PCRS score at Time 1 | $22.6 \pm 3.9$ |
| HR (bpm) at Time 1 | $72.9 \pm 10.2$ |
| lnvmHRV at Time 1 | $3.66 \pm 0.46$ |

Data are reported as means ± standard deviation. Abbreviations: BMI = body mass index; HR = heart rate; lnvmHRV = natural logarithm of vagally-mediated heart rate variability; ERQ = emotion regulation questionnaire; CESD = Center for Epidemiological Studies Depression Scale; PCRS = Perceived Coronavirus Risk scale.

**Table 2.** Linear regressions for the prediction of scores on the cognitive reappraisal and emotional suppression subscales of the ERQ based on vmHRV values at Time 0 and months elapsed between Time 1 and Time 0.

| **Dependent: ERQ reappraisal** |  |  |  |  |  |  |
| --- | --- | --- | --- | --- | --- | --- |
|  | *Unstandardized B* | *SE* | *t* | *p* | 95% *CI*<br>Lower<br>Bound | 95% *CI*<br>Upper<br>Bound |
| Model 1 |  |  |  |  |  |  |
| (Constant) | 15.09 | 2.06 | 7.33 | < .0001 | 11.05 | 19.13 |
| lnvmHRV | 3.42 | .54 | 6.37 | < .0001 | 2.36 | 4.47 |
| Model 2 |  |  |  |  |  |  |
| (Constant) | 20.03 | 3.04 | 6.59 | < .0001 | 14.07 | 25.99 |
| lnvmHRV | 2.99 | .57 | 5.27 | < .0001 | 1.88 | 4.11 |
| Months | -.16 | .07 | -2.20 | .028 | -.29 | -.017 |
| **Dependent: ERQ suppression** |  |  |  |  |  |  |
|  | *Unstandardized B* | *SE* | *t* | *p* | 95% *CI*<br>Lower<br>Bound | 95% *CI*<br>Upper<br>Bound |
| Model 1 |  |  |  |  |  |  |
| (Constant) | 16.55 | 1.08 | 15.37 | < .0001 | 14.44 | 18.66 |
| lnvmHRV | -.69 | .281 | -2.44 | .015 | -1.24 | -.13 |
| Model 2 |  |  |  |  |  |  |
| (Constant) | 20.61 | 1.58 | 13.05 | < .0001 | 17.50 | 23.71 |
| lnvmHRV | -1.03 | .29 | -3.49 | .001 | -1.61 | -.45 |
| Months | -.13 | .04 | -3.48 | .001 | -.20 | -.04 |

Abbreviations: lnvmHRV = natural logarithm of vagally-mediated heart rate variability; ERQ = emotion regulation questionnaire; *SE* = standard error; *CI* = confidence interval.

**Table 3.** Random effects regression models for the prediction of momentary levels of feeling “worried” and “safe” at Time 1 based on vmHRV values at Time 0 and months elapsed between Time 1 and Time 0.

| **Dependent: Worried** |  |  |  |  |  |  |  |
| --- | --- | --- | --- | --- | --- | --- | --- |
|  | Estimate | *SE* | *df* | *t* | *p* | 95% *CI*<br>Lower<br>Bound | 95% *CI*<br>Upper<br>Bound |
| Intercept | 6.67 | 1.79 | 63.82 | 3.73 | < .0001 | 3.09 | 10.24 |
| lnvmHRV | -1.27 | .42 | 63.03 | -3.01 | .004 | -2.12 | -.43 |
| Months | -.21 | .08 | 63.86 | -2.67 | .010 | -.37 | -.05 |
| lnvmHRV*Months | .04 | .019 | 63.27 | 2.29 | .025 | -.01 | -.08 |
| **Dependent: Safe** |  |  |  |  |  |  |  |
|  | Estimate | *SE* | *df* | *t* | *p* | 95% *CI*<br>Lower<br>Bound | 95% *CI*<br>Upper<br>Bound |
| Intercept | -1.20 | 2.25 | 66.37 | -.53 | -.534 | -5.70 | 3.29 |
| lnvvmHRV | 1.20 | .54 | 65.98 | 2.25 | .028 | .14 | 2.27 |
| Months | .08 | .10 | 66.37 | .77 | .442 | -.12 | .28 |
| lnvmHRV*Months | -.04 | .02 | 66.08 | -1.77 | .082 | -.09 | .01 |

Abbreviations: lnvmHRV = natural logarithm of vagally-mediated heart rate variability; *SE* = standard error; *CI* = confidence interval.

Partial correlation analyses (controlling for age and sex) revealed that vmHRV at T1 was positively associated with vmHRV at T0 (*r* = .631, *p* < .001) and negatively associated with CESD (*r* = -.261, *p* = .050) and ERQ suppression (*r* = -.255, *p* = .055) scores. Importantly, a positive significant correlation was found between CESD and PCRS scores (*r* = .391, *p* < .001). No significant correlations were found between vmHRV at T1 and PCRS (*r* = -.210, *p* = .117) and ERQ reappraisal (*r* = .147, *p* = .117) scores.

## DISCUSSION

The major and novel findings of the current study is that higher tonic HRV - assessed almost 2 years before - predicted a stronger engagement in more functional emotion regulation strategies, as well as higher daily feelings of safeness and reduced daily worry during COVID-19-related forced lockdown. These longitudinal findings strengthen previous cross-sectional studies showing that resting vmHRV is negatively correlated with dysfunctional emotion regulation strategies, such as the inability to accept negative emotions and subsequent emotional suppression (25), and positively correlated with more functional strategies, such as cognitive reappraisal (26, 27), acceptance (27, 28), self-compassion (29), and overall accessibility to emotion regulation strategies (30, 31). Of note, a negative correlation between vmHRV and emotional suppression strategies was also found in the present investigation. These results are particularly relevant in the context of stressful situations such as the COVID-19 pandemic, considering that maladaptive emotion regulation has been associated with negative psychological and somatic health outcomes (32). Relatedly, we found that self-reported depressive symptoms over the forced lockdown were positively correlated with the subjective perception of COVID-19 risk and negatively correlated with concurrent measures of vmHRV. Remarkably, in line with the neurovisceral integration theoretical perspective (10), higher pre-COVID-19 tonic vmHRV levels predicted reduced worrisome affective state and increased sense of safeness experienced for two consecutive days over the course of the lockdown, suggesting that HRV might be not only be a marker of but also a precursor of vulnerability/resilience to stress. Notably, momentary levels of sadness, anxiety and optimism were not significantly predicted by HRV at T0, pointing to the specificity of this biomarker for the inner sense of safeness. This is in line with the idea that HRV measures index the activity of a core set of neural structures that continuously assess the environment for signs of threat and safety and prepare the organism to adaptively regulate cognition, behavior, and physiology (33).

In interpreting our findings, we must acknowledge several limitations. First, due to difficulties in recruiting participants with reliable measures of vmHRV before the pandemic, the sample size is modest and should be extended in the future. Second, we obtained vmHRV measures at T1 with the use of a smartphone app, which could be suboptimal because smartphone apps acquire data using photoplethysmography and cannot discriminate between sinus and non-sinus beats. Nevertheless, we found a significant correlation between vmHRV measures at T1 and T0. Lastly, we do not have measures of emotional regulation strategies at T0, which would have allowed to obtain stronger evidence of the role of HRV as a predictor of coping styles during stressful situations. Notwithstanding these limitations, the present results support the implementation of HRV assessment to identify individuals who are at higher risk to suffer from stress-related symptoms (34) and encourage the use of early interventions such as transcutaneous vagus nerve stimulation, HRV biofeedback and compassion focused interventions to increase vmHRV as a way to foster resilience during challenging pandemic periods and beyond (35, 36).

## Data Availability

Data are available upon request to the authors.

## Notes

### Competing Interest Statement

The authors have declared no competing interest.

### Funding Statement

No funding to declare.

### Author Declarations

IRBs of Kings College London LRS-19/20-18429:COVID-19 and Sapienza University of Rome Prot. 0000653

## References

1. Organization WH. WHO Director-General’s opening remarks at the media briefing on COVID-19. 2020.

2. Dubey S, Biswas P, Ghosh R, et al. Psychosocial impact of COVID-19. Diabetes & metabolic syndrome. 2020; 14:779–788.

3. Rajkumar RP. COVID-19 and mental health: A review of the existing literature. Asian Journal of Psychiatry. 2020; 52:102066.

4. Rubin GJ, Wessely S. The psychological effects of quarantining a city. Bmj. 2020; 368:m313.

5. Bao Y, Sun Y, Meng S, Shi J, Lu L. 2019-nCoV epidemic: address mental health care to empower society. The Lancet. 2020; 395:e37–e38.

6. Corpuz JCG. COVID-19 and Mental Health. J Psychosoc Nurs Ment Health Serv. 2020; 58:4.

7. Holmes EA, O’Connor RC, Perry VH, et al. Multidisciplinary research priorities for the COVID-19 pandemic: a call for action for mental health science. The Lancet Psychiatry. 2020; 7(6):547–560

8. Porges SW. The polyvagal theory: new insights into adaptive reactions of the autonomic nervous system. Cleveland Clinic journal of medicine. 2009; 76 Suppl 2:S86–S90.

9. Carnevali L, Thayer JF, Brosschot JF, Ottaviani C. Heart rate variability mediates the link between rumination and depressive symptoms: A longitudinal study. International Journal of Psychophysiology. 2018; 131:131–138.

10. Thayer JF, Lane RD. A model of neurovisceral integration in emotion regulation and dysregulation. Journal of affective disorders. 2000; 61:201–216.

11. Lee SM, Han H, Jang K-I, et al. Heart rate variability associated with posttraumatic stress disorder in victims’ families of sewol ferry disaster. Psychiatry research. 2018; 259:277–282.

12. Bourdillon N, Yazdani S, Schmitt L, Millet GP. Effects of COVID-19 lockdown on heart rate variability. PloS one. 2020; 15:e0242303.

13. Cabrera A, Kolacz J, Pailhez G, Bulbena-Cabre A, Bulbena A, & Porges SW. Assessing body awareness and autonomic reactivity: Factor structure and psychometric properties of the Body Perception Questionnaire-Short Form (BPQ-SF). International journal of methods in psychiatric research. 2018; 27(2), e1596.

14. Kolacz J, Dale LP, Nix EJ, et al. Adversity History Predicts Self-Reported Autonomic Reactivity and Mental Health in US Residents During the COVID-19 Pandemic. Frontiers in psychiatry. 2020; 11:577728–577728.

15. Miller JG, Chahal R, Kirshenbaum JS, et al. Heart Rate Variability Moderates the Link Between COVID-19 Stress and Emotional Problems in Adolescents: Evidence for Differential Susceptibility. 2020; 1–12.

16. Di Bello M, Ottaviani C, Petrocchi N. Compassion is not a benzo: Distinctive associations of Heart Rate Variability with its empathic and action components. Frontiers in Neuroscience. 2021; 15:223.

17. Carnevali, Pattini E, Sgoifo A, Ottaviani C. Effects of prefrontal transcranial direct current stimulation on autonomic and neuroendocrine responses to psychosocial stress in healthy humans. Stress. 2020; 23:26–36.

18. Sgoifo A, Carnevali L, Pattini E, et al. Psychobiological evidence of the stress resilience fostering properties of a cosmetic routine. Stress. 2021; 24:53–63.

19. Hohenschurz-Schmidt DJ, Calcagnini G, Dipasquale O, et al. Linking pain sensation to the autonomic nervous system: the role of the anterior cingulate and periaqueductal gray resting-state networks. Frontiers in Neuroscience. 2020; 14:147.

20. Kanovsky M, Halamová J. Perceived Threat of the Coronavirus and the Role of Trust in Safeguards: A Case Study in Slovakia. Frontiers in psychology. 2020; 11:3371.

21. Radloff LS. The CES-D scale: A self-report depression scale for research in the general population. Applied psychological measurement. 1977; 1:385–401.

22. Spaapen DL, Waters F, Brummer L, Stopa L, Bucks RS. The emotion regulation questionnaire: validation of the ERQ-9 in two community samples. Psychological Assessment. 2014; 26:46.

23. Childs A. Qualtrics. 2020.

24. Laborde S, Mosley E, Thayer JF. Heart rate variability and cardiac vagal tone in psychophysiological research–recommendations for experiment planning, data analysis, and data reporting. Frontiers in psychology. 2017; 8:213.

25. Visted E, Sørensen L, Osnes B, et al. The association between self-reported difficulties in emotion regulation and heart rate variability: the salient role of not accepting negative emotions. Frontiers in psychology. 2017; 8:328.

26. Denson TF, Grisham JR, Moulds ML. Cognitive reappraisal increases heart rate variability in response to an anger provocation. Motivation and Emotion. 2011; 35:14–22.

27. Gray JM, Tully EC. Cognitive reappraisal moderates the quadratic association between heart rate variability and negative affectivity. Psychophysiology. 2020; 57:e13584.

28. O’Connor M-F, Allen JJ, Kaszniak AW. Autonomic and emotion regulation in bereavement and depression. Journal of psychosomatic research. 2002; 52:183–185.

29. Svendsen JL, Osnes B, Binder P-E, et al. Trait self-compassion reflects emotional flexibility through an association with high vagally mediated heart rate variability. Mindfulness. 2016; 7:1103–1113.

30. Gillie BL, Vasey MW, Thayer JF. Heart rate variability predicts control over memory retrieval. Psychological Science. 2014; 25:458–465.

31. Aldao A, Mennin DS. Paradoxical cardiovascular effects of implementing adaptive emotion regulation strategies in generalized anxiety disorder. Behaviour research and therapy. 2012; 50:122–130.

32. Hu T, Zhang D, Wang J, et al. Relation between emotion regulation and mental health: a meta-analysis review. Psychological reports. 2014; 114:341–362.

33. Thayer JF, Åhs F, Fredrikson M, Sollers III JJ, Wager TD. A meta-analysis of heart rate variability and neuroimaging studies: implications for heart rate variability as a marker of stress and health. Neuroscience & Biobehavioral Reviews. 2012; 36:747–756.

34. Thayer JF, Drury RL. Heart Rate Variability in Assessing Community COVID-19. Frontiers in Neuroscience. 2021; 15:646.

35. Dedoncker J, Vanderhasselt M-A, Ottaviani C, Slavich GM. Mental health during the COVID-19 pandemic and beyond: The importance of the vagus nerve for biopsychosocial resilience. Neuroscience & Biobehavioral Reviews. 2021.

36. Steffen PR, Foxx J, Cattani K, et al. Impact of a 12-week group-based compassion focused therapy intervention on heart rate variability. Applied Psychophysiology and Biofeedback. 2021; 46:61–68.

